# Racial/Ethnic Disparities in Hospital Admissions from COVID-19 and Determining the Impact of Neighborhood Deprivation and Primary Language

**DOI:** 10.1101/2020.09.02.20185983

**Authors:** Nicholas E. Ingraham, Laura N. Purcell, Basil S. Karam, R. Adams Dudley, Michael G. Usher, Christopher A. Warlick, Michele L. Allen, Genevieve B. Melton, Anthony Charles, Christopher J. Tignanelli

## Abstract

**Background:** Despite past and ongoing efforts to achieve health equity in the United States, persistent disparities in socioeconomic status along with multilevel racism maintain disparate outcomes and appear to be amplified by COVID-19.

**Objective:** Measure socioeconomic factors and primary language effects on the risk of COVID-19 severity across and within racial/ethnic groups.

**Design:** Retrospective cohort study.

**Setting:** Health records of 12 Midwest hospitals and 60 clinics in the U.S. between March 4, 2020 to August 19, 2020.

**Patients:** PCR+ COVID-19 patients.

**Exposures:** Main exposures included race/ethnicity, area deprivation index (ADI), and primary language.

**Main Outcomes and Measures:** The primary outcome was COVID-19 severity using hospitalization within 45 days of diagnosis. Logistic and competing-risk regression models (censored at 45 days and accounting for the competing risk of death prior to hospitalization) assessed the effects of neighborhood-level deprivation (using the ADI) and primary language. Within race effects of ADI and primary language were measured using logistic regression.

**Results:** 5,577 COVID-19 patients were included, 866 (n=15.5%) were hospitalized within 45 days of diagnosis. Hospitalized patients were older (60.9 vs. 40.4 years, p<0.001) and more likely to be male (n=425 [49.1%] vs. 2,049 [43.5%], p=0.002). Of those requiring hospitalization, 43.9% (n=381), 19.9% (n=172), 18.6% (n=161), and 11.8% (n=102) were White, Black, Asian, and Hispanic, respectively.

Independent of ADI, minority race/ethnicity was associated with COVID-19 severity; Hispanic patients (OR 3.8, 95% CI 2.72–5.30), Asians (OR 2.39, 95% CI 1.74–3.29), and Blacks (OR 1.50, 95% CI 1.15–1.94). ADI was not associated with hospitalization. Non-English speaking (OR 1.91, 95% CI 1.51–2.43) significantly increased odds of hospital admission across and within minority groups.

**Conclusions:** Minority populations have increased odds of severe COVID-19 independent of neighborhood deprivation, a commonly suspected driver of disparate outcomes. Non-English-speaking accounts for differences across and within minority populations. These results support the continued concern that racism contributes to disparities during COVID-19 while also highlighting the underappreciated role primary language plays in COVID-19 severity across and within minority groups.

**Key Points:** *Question:* Does socioeconomic factors or primary language account for racial disparities in COVID-19 disease severity?

*Findings:* In this observational study of 5,577 adults, race/ethnicity minorities and non-English as a primary language, independent of neighborhood-level deprivation, are associated with increased risk of severe COVID-19 disease.

*Meaning:* Socioeconomic factors do not account for racial/ethnic disparities related to COVID-19 severity which supports further investigation into the racism and highlights the need to focus on our non-English speaking populations.

## Introduction

Health inequity during COVID-19 has been documented through numerous studies since it was brought to light through the landmark study of the five boroughs in New York City. The study found the Bronx to have the highest hospitalization and death rate of COVID-19.(1) The Bronx also happens to be the borough with the largest non-White racial/ethnic minority populations, lowest income, and lowest education level.(1) Studies in the United Kingdom found lower socioeconomic status (SES) and minority patients are more likely to be infected with COVID-19 even after adjusting for risk behaviors.(2) Ecological studies found areas with high poverty and predominantly minority populations have infection rates 8 times higher than high poverty areas with predominantly White patients.(3) As noted in the paper, community-level analysis, without controlling for individual-level differences, limits the ability to account for individual factors which would strengthen the concerns that this association may, in fact be causal. Furthermore, language barriers have been associated with outcomes but limited data regarding its impact in COVID-19, as it relates to racial disparities, currently exist.(4-6)

Racial and ethnic disparities in health are well documented in the United States and their elimination is an important public health and societal imperative.(7, 8) The etiology of health disparities is multifactorial, especially with racism’s multi-level societal construct which plays a key role in the challenges we face along the path to health equity.(9-11) Bailey et al. highlight one of the foundational levels, structural bias.(9) Structural racism, which contributes to residential segregation, discriminatory incarceration, and inequitable access to healthcare services, is an integral factor contributing to health disparities. Due to structural racism and the resulting disparate influences of social determinants of health, along with poor access to healthcare, racial and ethnic minorities have higher rates of chronic comorbidities leading to increased susceptibility to critical illnesses.(12-14) While many of these concerning disparities can be traced back to structural biases with healthcare access, income, and residential segregation, it is important to note that institutional racism exists through sometimes subtle and unassuming policies (e.g., family visitation restrictions)(15) that exist despite contrary evidence.(16) These examples highlight the complexity and intertwined factors contributing to disparate outcomes for the critically ill. The complex nature of the issue mandates an equally complex, yet appropriately designed, effort to adequately address this public health crisis.

COVID-19 has created a unique research environment by bringing together public health, clinical, and epidemiological researchers at an unprecedented level.(17) The amount of granular data being collected along with newly founded collaborations provides an opportunity to study healthcare disparities through the lens of the pandemic. Few studies have accounted for the much needed assessment that takes into account socioeconomic variables along with granular patient characteristics to appropriately tease out the different drivers.(18) We hypothesize that neighborhood-level deprivation and/or primary language spoken may account for the disproportionate outcomes seen in racial/ethnic minorities. Our aim was to examine how neighborhood deprivation, primary language, and race/ethnicity effect the risk of COVID-19 severity. Having a better understanding of these complex relationships are critical to guiding the ongoing prevention, policy, and intervention efforts aimed at fighting this global pandemic in an equitable fashion.(18)

## Methods

### Design and Data

Retrospective analysis of 12 Midwest hospitals and 60 primary care clinics, between March 4 - August 19, 2020. To account for patient transfers across hospitals or clinics data was pooled across different electronic health records (EHRs) and a unique patient identifier was created accounting for clinic, ED, or hospital encounters across systems. In cases where a patient was seen in two different EHR systems, the most recent EHR comorbidity records were utilized. All patients that opted out of research were excluded from analysis. The COVID-19 datamart includes individual-level data for individuals with PCR-confirmed COVID-19, covering a diverse range of ages, races, ethnicities, and geographic regions within the Midwest. This study was approved by the University of Minnesota institutional review board (STUDY00001489) which provided a waiver of consent for this study.

### Population

Included in this study were individuals with PCR-confirmed COVID-19 that did not opt out of research (<2.5% of patients opt out of research). Patients with missing race or ethnicity data were excluded from this analysis (1,225 patients, 17.9%) and patients that developed new onset COVID-19 following hospital admission for another reason (13 patients).

### Outcomes

Our primary outcome was the need for hospitalization within 45 days of PCR confirmed COVID-19. Individuals not hospitalized by August 24, 2020 were censored in the competing risks regression model and treated as not requiring hospitalization in the logistic regression model.

### Independent Variable and Confounding Variables

The independent variables were race/ethnicity (White, Black, Asian, Other) or ethnicity (Hispanic or non-Hispanic), English vs non-English speaking, and the National Area Deprivation Index (ADI) categorized into quintiles (1^st^ quintile = lowest neighborhood-level of deprivation, 5^th^ = highest neighborhood-level of deprivation). Race/ethnicity was collected for administrative purposes and quality was not verified. The Area Deprivation Index’s development and utility has been described previously.(19) In brief, the ADI is a measure of the socioeconomic disadvantage of a neighborhood (using the 9 digit zip code) calculated from 17 different indicators that includes education, housing (e.g. occupancy rates, household value), poverty (e.g. median income, number of dependents, % with complete plumbing), and employment status. ADI(20) and other indices(21-23) have been used to estimate the effects of these socioeconomic variables in aggregate on outcomes, including in COVID-19.

In addition to the independent variables, models were adjusted for age, gender, Elixhauser Comorbidity Index,(24) relationship status, and rurality/urbanity. Urbanity was defined as living in a zip code with a population density > 200 people per square mile.(25) Marital status was characterized as single, married (domestic partner, life partner, or married), divorced or legally separated, or widowed.

### Statistical Analysis

Continuous variables with a skewed distribution were expressed by median and interquartile range (IQR). Categorical variables were expressed by percentages. Univariate analysis compared those admitted to those without admission. The association of race/ethnicity on mortality, hospital admission severity (laboratory values and vitals), in-hospital complications (cardiovascular, respiratory, hematologic, renal, infectious), ICU and mechanical ventilation utilization and in-hospital mortality was evaluated for patients requiring hospital admission. Overall missingness was low (0 - 2.06%). Given the low rate of missingness, imputation was deemed unnecessary.(26)

Logistic and competing-risk regression models (censored at 45 days while accounting for the competing risk of death prior to hospitalization) were used to assess the independent association of race/ethnicity, ADI, and primary language (English vs other) on the primary outcome. Comorbidities were determined using a random forest variable importance analysis and models were compared using the comorbidities selected by variable importance analysis (Appendix Table 1) to aggregate indices of comorbidities commonly used to adjust for baseline conditions (Elixhauser) given the limited experience of adjusting for co-morbid conditions in COVID-19. The final model to be determined by the best Bayesian Information Criterion (BIC).

We investigated the different effects of ADI and primary language across and between race/ethnic groups and the association with risk for admission. By plotting the cumulative incidence of hospital admission it was clear there were consistent and sequential increases across and within groups in relation to both ADI and primary language. To estimate if these differences within race/ethnic groups were significant, logistic regression analysis (using the same covariates as the main model) were performed by stratifying either ADI or primary language by race. Each model used the race of interest as the base to determine if ADI or language differed within each race. For ADI models, we report the comparison between the 5 ^th^ quintile (highest level) to the 1^st^ quintile (lowest level of area deprivation).

Statistical analyses were performed using Stata MP, version 16 (StataCorp, College Station, TX). Statistical significance was defined as a two-tailed p-value < 0.05.

## Results

Of the 5,577 COVID-19 patients meeting inclusion criteria for this study, 866 (n=15.5%) patients were admitted to the hospital within 45 days of COVID-19 diagnosis.(Figure 1) In the overall COVID-19 cohort, the median age was 43.7 years [IQR: 27.4 - 62.3] and 44.4% (n=2,474) were male. Patients were predominantly White (n=2,931, 52.6 %), followed by Black (n=1,225, 22.0%), Asian (n=677, 12.2%), and Hispanic (n=416, 7.5%). The median national ADI was 37% (IQR: 22-48%).

**Figure 1:**
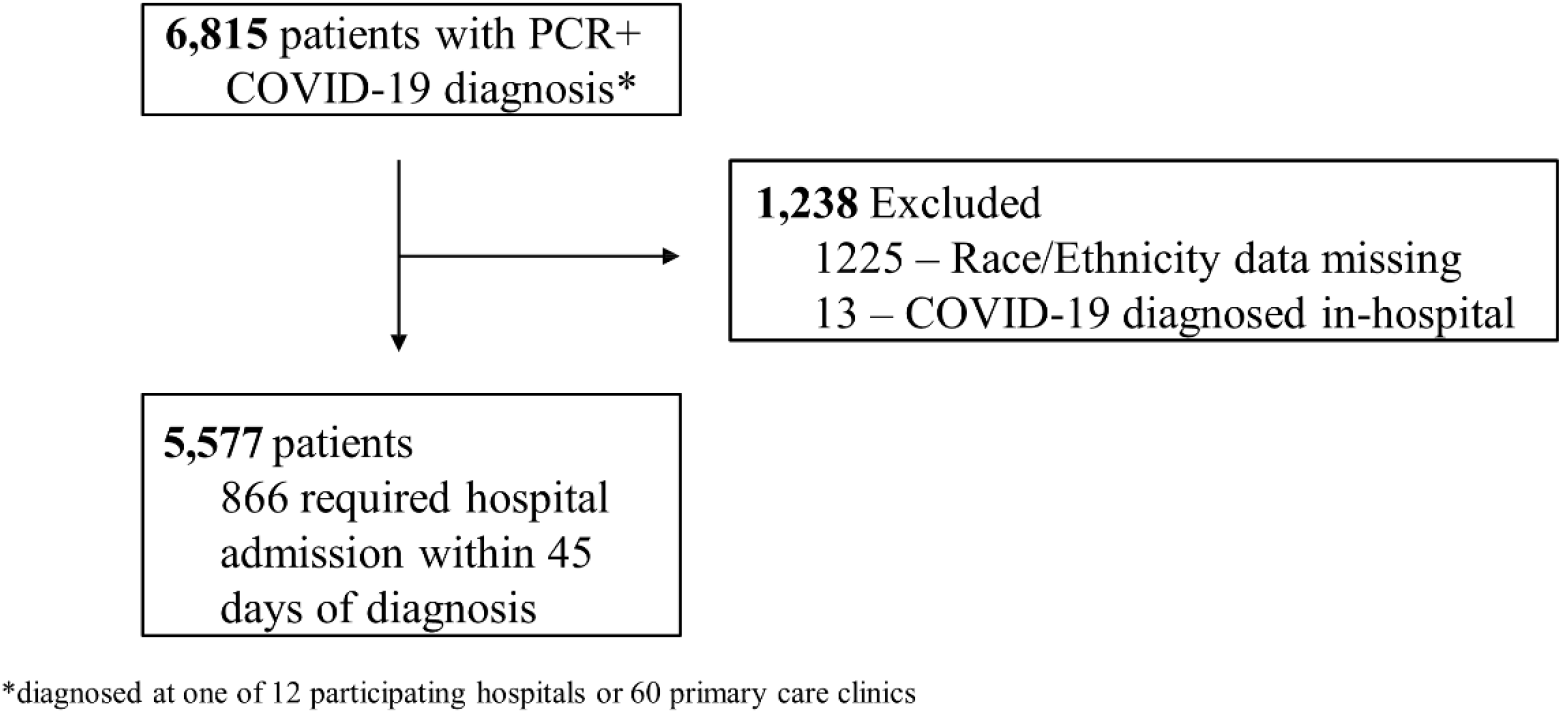
Inclusion / Exclusion Diagram Study diagram of patients included in final analysis of COVID-19 Data Registry

Patients who required hospitalization within 45 days of diagnosis, a marker of severe disease, were older (60.9 years [IQR: 45.7-75.9] vs. 40.4 year [IQR: 25.6-58.3], p<0.001) and a greater proportion were male (n=425, 49.1% vs. n=2,049, 43.5%, p=0.002). Table 1. More of the admitted versus non-admitted cohort live in fourth and fifth ADI quartile neighborhoods. A higher proportion of non-English speaking patients required admission (n=301, 34.8% vs. n=785, 16.7%, p<0.001). Patients who were admitted had higher Elixhauser Comorbidity Score (5.0, IQR: 3.0-9.0 vs. 1.0, IQR: 0.0-2.0, p<0.001) and had a higher mortality (n=108, 12.5% vs. n=68, 1.4%, p<0.001) than those not requiring admission.

**Table 1:** Univariate analysis of COVID-19 patients not admitted compared to those admitted to the hospital. Univariate analysis comparing PCR+ C0VID-19 patients who were admitted within 45 days of testing vs those without hospital admission. ADI quintiles represent lowest areas of deprivation (1st quintile) to the highest areas of deprivation (5th quintile).

|  | Not Admitted<br>(n=4,711, 84.5%) | Admitted<br>(n=866, 15.5%) | p-value |
| --- | --- | --- | --- |
| **Age (years):** median (IQR) | 40.4 (25.6-58.3) | 60.9 (45.7-75.9) | <0.001 |
| **Male Sex:** n (%) | 2,049 (43.5) | 425 (49.1) | 0.002 |
| **Race/ethnicity:** n (%) |  |  |  |
| White | 2,550 (54.1) | 381 (44.0) | <0.001 |
| Black | 1,053 (22.4) | 172 (19.9) |  |
| Asian | 516 (11.0) | 161 (18.6) |  |
| Hispanic | 314 (6.7) | 102 (11.8) |  |
| Declined | 219 (4.6) | 32 (3.7) |  |
| Other | 59 (1.3) | 18 (2.1) |  |
| **Non-English Speaking:** n (%) | 785 (16.7) | 301 (34.8) | <0.001 |
| **Area Deprivation Quintiles:** n (%) |  |  |  |
| First: 0-20% | 1,005 (21.3) | 169 (19.5) | <0.001 |
| Second: 21-40% | 1,559 (33.1) | 237 (27.4) |  |
| Third: 41-60% | 1,155 (24.5) | 220 (25.4) |  |
| Fourth: 61-80% | 657 (13.9) | 142 (16.4) |  |
| Fifth: 81-100% | 335 (7.1) | 98 (11.3) |  |
| **Urban:** n (%) | 3,416 (85.8) | 728 (93.7) | <0.001 |
| **Marital Status:** n (%) |  |  |  |
| Single | 2,428 (51.5) | 324 (37.4) | <0.001 |
| Married | 1,837 (39.0) | 381 (44.0) |  |
| Separated/Divorced | 236 (5.0) | 73 (8.4) |  |
| Widowed | 210 (4.5) | 88 (10.2) |  |
| **Comorbidities:** n (%) |  |  |  |
| Hypercoagulable State | 39 (0.8) | 33 (3.8) | <0.001 |
| Hypocoagulable State | 211 (4.5) | 242 (27.9) | <0.001 |
| Type 1 Diabetes Mellitus | 95 (2.0) | 84 (9.7) | <0.001 |
| Type 2 Diabetes Mellitus | 578 (12.4) | 338 (39.0) | <0.001 |
| Atrial Fibrillation or Atrial Flutter | 220 (4.7) | 163 (18.8) | <0.001 |
| Hypertension | 1,333 (28.6) | 574 (66.3) | <0.001 |
| Chronic Kidney Disease | 334 (7.2) | 241 (27.8) | <0.001 |
| Chronic Obstructive Pulmonary Disease | 169 (3.6) | 123 (14.2) | <0.001 |
| Liver Disease | 237 (5.1) | 131 (15.1) | <0.001 |
| Cerebral Vascular Disease | 279 (6.0) | 151 (17.4) | <0.001 |
| Sleep Apnea | 308 (6.6) | 154 (17.8) | <0.001 |
| Congestive Heart Failure | 246 (5.3) | 174 (20.1) | <0.001 |
| Obesity | 849 (18.2) | 294 (33.9) | <0.001 |
| **Mortality: n (%)** | **68 (1.4)** | **108 (12.5)** | **<0.001** |

Of those requiring hospitalization, 43.9% (n=381), 19.9% (n=172), 18.6% (n=161), and 11.8% (n=102) were White, Black, Asian, and Hispanic, respectively. White patients (69.6 years, IQR 55.4-82.2) were significantly older than their Black (55.4 years, IQR 38.2-70.8), Asian (58.9 years, IQR 41.8-69.5), and Hispanic (48.5 years, IQR 40.4-56.7) counterparts, p<0.001. Approximately half of White and Hispanic patients lived in the first and second quintile ADI neighborhoods, in contrast, nearly the same proportion of Asian patients lived in the fourth quintile, and Black patients were evenly distributed over all ADI quintiles. White patients (6.0, IQR 4.0-10.0) had the highest and Hispanic patients (3.5, IQR 1.0-6.0) had the lowest Elixhauser Comorbidity scores, p<0.001 of those admitted to the hospital. White patients (n=58, 15.2%) had a higher in-hospital mortality than the Black (n=12, 7.0%), Asian (n=16, 9.9%), and Hispanic (n=4, 3.9%) cohorts, p<0.001. Admission labs were not significantly different between racial/ethnic groups, specifically D-dimer and IL-6 which have been shown to be associated with mortality risk on admission in COVID-19.(27, 28) Appendix Table 2.

When choosing our final model for analysis, calibration and discrimination were evaluated following each model using Hosmer Lemeshow goodness of fit (p > 0.2) and area under the receiver operating characteristic curve (AUROC) > 0.8 for all models. Logistic regression models with Elixhauser performed better (BIC 3686) compared to using selected co-morbidities (BIC 4008) as described above and thus the final model included Elixhauser in attempts to adjust for co-morbidities.

### Primary Analysis

On logistic regression analysis, minority race/ethnicity was an independent predictor for hospital admission. (Table 2) Hispanic patients (OR 3.8, 95% CI 2.72 - 5.30), Asians (OR 2.39, 95% CI 1.74 - 3.29, and Blacks (OR 1.50, 95% CI 1.15 - 1.94) had higher odds of hospital admission within 45 days compared to White patients. Surprisingly, ADI was not independently associated at any quintile, while being non-English speaking (OR 1.91, 95% CI 1.51 - 2.43) was independently associated with an increased odds of hospitalization when compared to English speaking. On sensitivity analysis, to account for the competing risk of death and censoring at 45 days; aside from marital status, these associations remained significant in the competing-risk model. (Table 2)

**Table 2:**
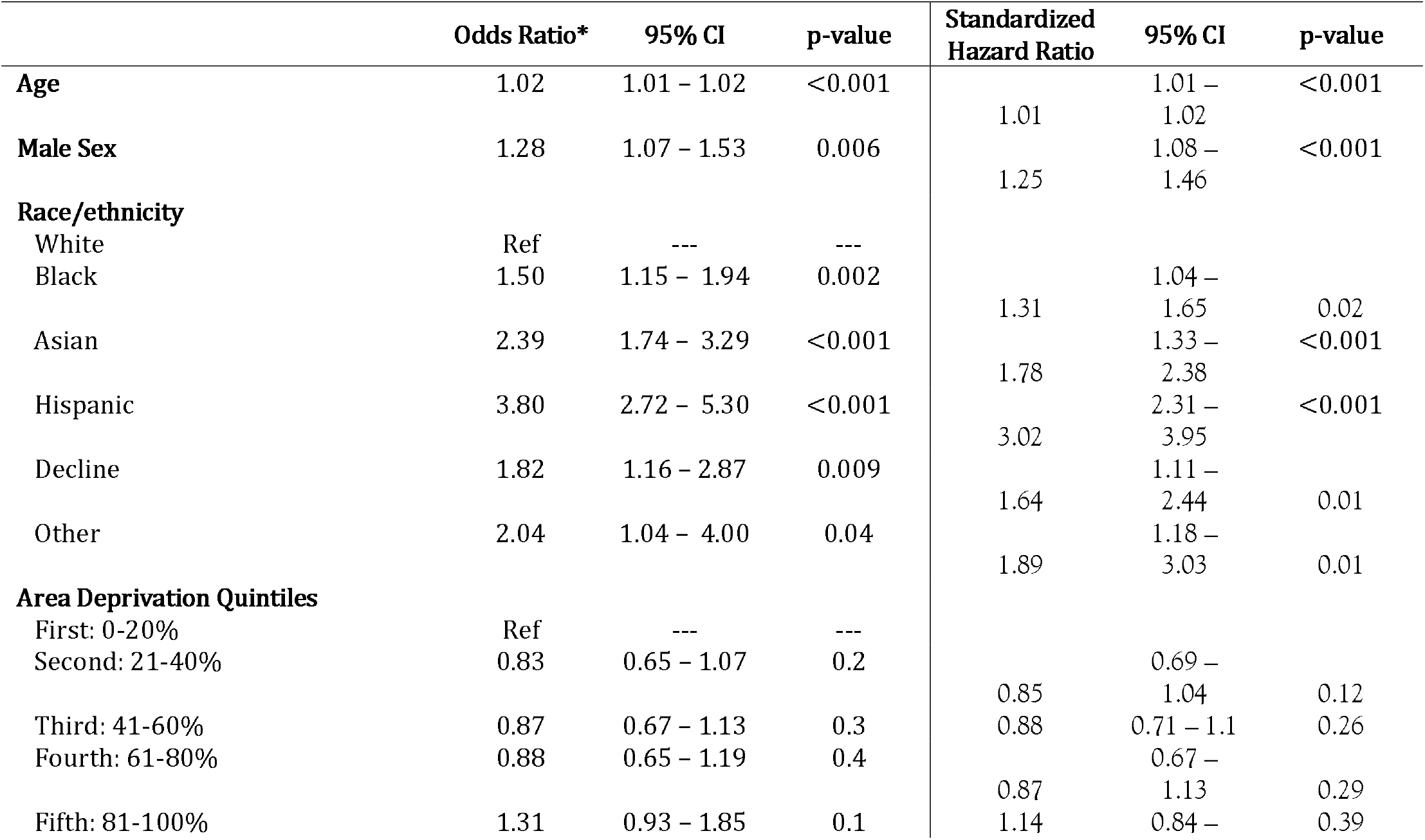
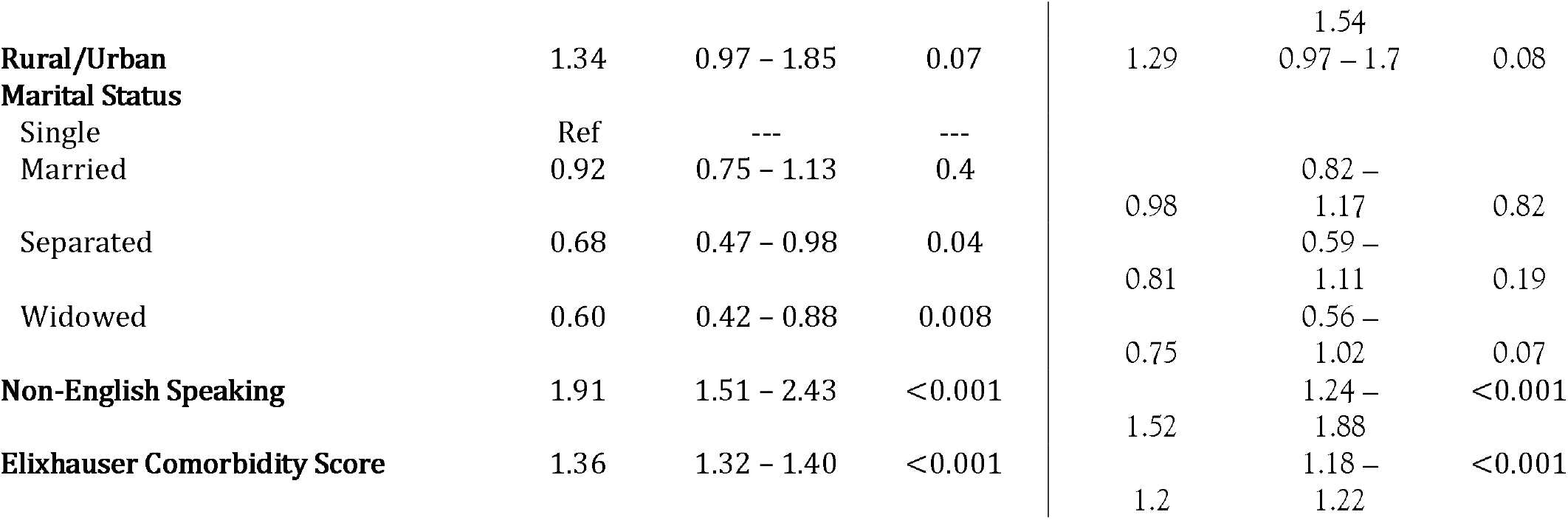
Multivariate logistic regression and competing-risk models for hospital admission in COVID-19 patients. Multivariate logistic regression (left) with odds of hospital admission in patients with PCR+ COVID-19 diagnosis within 45 days of testing. Competing risk model (right) with standardized hazard ratio of hospital admission in patients with PCR+ COVID-19 censored at 45 days from testing while accounting for death prior the primary endpoint. ADI quintiles represent lowest areas of deprivation (1^st^ quintile) to the highest areas of deprivation (5^th^ quintile). * AUROC: 0.854

### Race/Ethnicity ~ Area Deprivation Index

Cumulative incidence plots were generated to visualize the disparity by race/ethnicity and ADI quintile, (Figure 2a). On logistic regression, when compared to Whites living in the first ADI quintile, there is no statistical difference in hospitalization of Whites living in the fifth ADI quintile. In contrast, there is a sequential increase in odds of needing hospitalization of first (low neighborhood deprivation) ADI quintile Black patients (OR 1.75, 95% CI 1.05 - 2.93), fifth (high neighborhood deprivation) ADI quintile Black patients (OR 1.90, 95% CI 1.13 - 3.19), first quintile Asian patients (OR 2.31, 95% CI 1.02 - 5.26), fifth ADI quintile Asian (OR 4.85, 95% CI 1.86 - 12.60), and first ADI quintile Hispanic (OR 3.68, 95% CI 1.97 - 6.88), and fifth ADI quintile Hispanic (OR 10.14, 95% CI 3.85 - 26.74) when compared to White patients in the first ADI. (Appendix Table 2). On subgroup analysis comparing the fifth vs first quintile within each racial/ethnic group, there was no significant difference in the odds of hospitalization. (Figure 3a)

**Figure 2:**
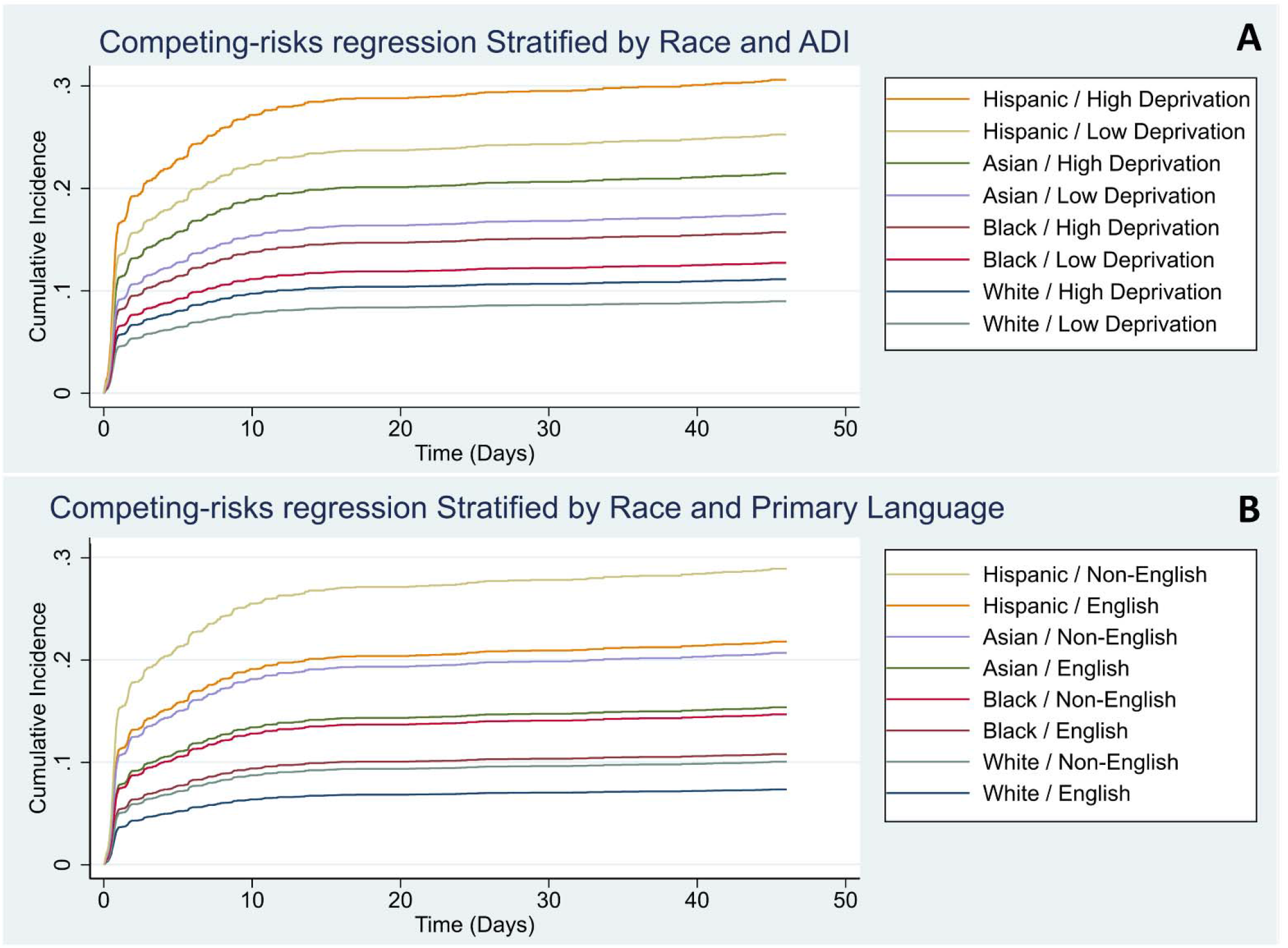
Competing risk regression cumulative incidence of hospital admission over time by ADI (A) and Primary Language (B) stratified by race/ethnicity. Competing risk regression cumulative incidence of hospital admission over time by ADI (A) and Primary Language (B) stratified by race/ethnicity. Models were censored at 45 days and accounted for death occurring prior to the primary endpoint (hospital admission).

**Figure 3:**
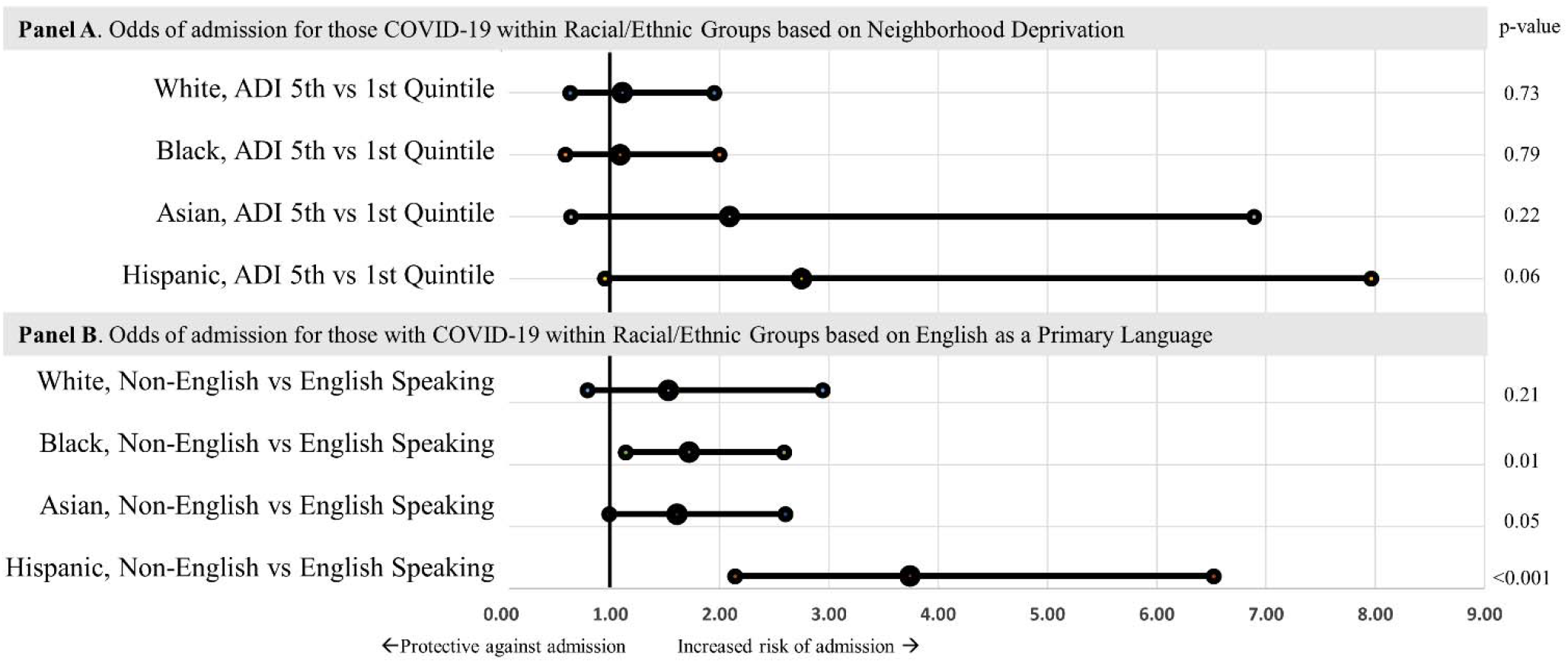
Forest Plots of multiple logistic regression models using each race/ethnicity as a baseline to compare ADI and Primary language stratified by race/ethnicity. Multivariate logistic regression models using each race/ethnicity as a baseline within its respective model to compare within each race/ethnicity high vs low neighborhood deprivation (top) and primary language (bottom) after adjusting for age, sex, marital status, urbanity, and comorbidities the odds of hospital admission in patients with PCR+ COVID-19 diagnosis within 45 days of testing.

### Race/Ethnicity ~ Primary Language

Cumulative incidence plots were generated to visualize the disparity by race/ethnicity and primary language (English vs non-English). (Figure 2b) Sequential increase in odds of hospitalization was similar to the above analysis with ADI when comparing each racial/ethnicity group by primary language (English or other) to English speaking Whites (data not shown). In contrast to the ADI models, on subgroup analysis, comparing those who speak English as their primary language vs. non-English speaking within each minority group, there was a significant difference in the odds of hospitalization; Black (OR 1.72. 95% CI 1.14-2.59), Asian (OR 1.61, 95% CI 1.0-2.6), and Hispanic (3.74, 95% CI 2.14-6.52). (Figure 3b)

## Discussion

In this study, we assessed outcomes of patients diagnosed with COVID-19 in the outpatient setting and investigated racial/ethnic difference and how neighborhood-level deprivation (ADI) and English as a primary language were associated with the need for hospital admission within 45 days (as a surrogate for COVID-19 severity. We demonstrated race/ethnicity was independently associated with hospital admission in COVID-19 positive patients, independent of area deprivation index which has been commonly thought to be a prominent driver of disparities during the pandemic. Furthermore, neighborhood-level deprivation was not associated with hospital admission but did have trends towards within race/ethnicity effects. Non-English speaking was associated with increased odds of admission, independent of race/ethnicity and neighborhood deprivation, overall and within minority subgroups. We also observed that after accounting for the competing risk of death these associations persisted. Importantly, despite our hypothesis that neighborhood deprivation and/or primary language may account for racial differences in outcomes, race/ethnicity remained statistically significant. These findings of a persistent association between race/ethnicity and COVID-19 severity support concerns of either racism or another unidentified confounder that is driving the association between skin color and COVID-19 severity.

Since the beginning of the pandemic, calls for urgent surveillance of this public health crisis and predictions of disparate outcomes that would emerge from COVID-19 were immediate.(29-31) These concerns were well founded, as it would be later demonstrated that Hispanic and Black patients have increased odds of hospital admission and high severity of illness due to COVID-19.(32, 33) Later studies have validated fears that socioeconomic factors are associated with COVID-19 and may account for at least some, if not a large proportion of racial disparities.(3, 34) To date, most studies using community-level analysis have uncovered numerous associations that suggest we are not only far from health equity, but COVID-19 is threatening to widen the gap.(35) For example, COVID-19 infection rates are higher in areas with a greater percentage of homes with overcrowding, lower education status, and limited access to healthcare.(34) Comorbidities, which are known to disproportionately affect racial minorities and impoverished populations, have been implicated in the association between worse outcomes of critical illness among these populations.(12-14) As observed with SARS-CoV,(36) comorbidities increase not only for the risk for infection but also severity of illness.(37, 38) Low health literacy was also predicted to contribute to health inequalities that may stem from the pandemic.(39) Survey studies regarding general disease knowledge and preventative strategies surrounding COVID-19 supported fears that lower health literacy could be associated with lack of disease understanding.(40) Compounding these issues further is the misinformation that accompanies this pandemic,(17) which also disproportionately affects individuals with lower education levels.(39) Attributing disparities in COVID-19 to socioeconomic variables or comorbidities in isolation would be inappropriate given the evidence that SES is associated with higher rates of comorbidities, subsequently complicating the causal diagram.(41)

Characterizing SES is complex in nature; however, the ADI accounts for key neighborhood-level characteristics. Area deprivation assesses poverty, housing, education, and employment. While it should not be used interchangeably with SES, it does incorporate a portion of socioeconomic factors that warrant assessment. Furthermore, ADI is able to account for some effects of environmental racism including the disproportionate degree of exposure to malignant environmental factors (i.e. pollution, food deserts) experienced by minority populations.(42) This could very well be driving some of the adverse health outcomes, and by using patients’ zip code to determine ADI, we sought to minimize the confounding of this variable. Limited data regarding area deprivation and COVID-19 exist, specifically in the United States. Studies in India(23) and the U.K.(21, 22) found areas of deprivation to be associated with increased risk of COVID-19. Both U.K. studies assessed COVID-19 patients and found that Black and Asian patients were at an increased odds of hospitalization. Similar to our findings, after controlling for areas of deprivation and comorbidities, the risk for racial and ethnic minority patients persisted.(21, 22) Furthermore, Hispanic patients had the highest odds of hospital admission despite having less comorbidities. These findings highlight and support the alarming fact that minorities have a higher risk of severe infection that is not attributable solely to baseline health conditions or socio-economic factors.

This contrasts with studies examining racial disparities in other disease processes. For example, it has been repeatedly shown that most of the racial disparities in outcomes of cancer patients can be explained by treatment received and socioeconomic status.(43-45) Similarly, in COPD patients, Black race was associated with higher disease severity but these differences disappeared after adjusting for SES and comorbid conditions.

The pandemic is putting a strain on normalcy for everyone; unfortunately, the largest burden of this strain appears to be on the most vulnerable which tips the scales further away from our goals of health equity. Evidence of racial differences seen with imaging in COVID-19 has been associated with language proficiency. Minority patients were found to have more severe lung edema determined by chest x-rays on admission.(4) Interestingly, part of the racial differences in imaging severity was mediated by English proficiency. General health studies have shown English proficiency may have a greater effect than health literacy.(5) Only one other study to date has found primary English language to be associated with COVID-19 outcomes, specifically, risk of infection.(6) Despite different outcomes in their study compared to ours (risk of infection vs risk of hospitalization, respectively) Rozenfeld et al., also found primary language other than English to be a greater risk factor than race itself. For these reasons we not only included primary language in our model, which notably was significantly associated with our outcome, but we further explored within race effects. Strikingly we found non-English speaking patients within each minority race/ethnic group to have higher odds of hospitalization compared to those within the same group but spoke English as their primary language.

Despite our best efforts to address potential racial and ethnic confounders, multiple limitations need to be considered when interpreting our results. First, the need for hospitalization may vary by site. With our current dataset, we are unable to determine whether that is the case for the population under study. Also, using hospitalization as an endpoint may be underwhelming (relative to other endpoints such as intensive care unit admission and mortality); however, there are many confounding factors that occur once the patient is hospitalized that may influence the observed effects. Hospitalization is a proxy for not only severity but also objective healthcare resource use and thus we argue, is not only of critical importance for physicians but can also assist administrators when assessing resource utilization in their communities. Testing deficiencies in low risk minorities (creating the perception of higher risk) has the potential to bias the results; however, the lack of testing in these patients is unlikely to alter the findings in our study and would only highlight another potential driver in observed disparities (i.e. lack of testing equality in the outpatient setting). Insurance status was not included in our analysis. However, this variable is commonly used to control for SES, and given that we have adjusted for multiple socioeconomic attributes, we feel this may attenuate the contribution insurance may have on our outcomes. Furthermore, there remains a possibility of residual confounding even after adjusting for SES, due to categorization of socioeconomic status variables, measurement error in socioeconomic indicators, and incommensurate socioeconomic indicators.(46) Finally, neighborhood-level SES (ADI) and individual level SES are sometimes discordant and attributing group level measures to individuals may limit interpretation.(47) However, in the absence of individual level socioeconomic attributes, reliance on aggregate measures is necessary to examine these racial disparities.

## Conclusion

Despite its success in many arenas and relative prosperity, paradoxically health equity persistently remains an aspiration in the United States. Studies like this and others are needed to continue taking steps forward. Failure to widely acknowledge, care for, and/or act upon these gaps makes the culprits that harder to tease out and increases the complexity of the intervention. Our findings highlight areas of neighborhood-level deprivation may contribute to racial disparities, but to a lesser degree when controlling individual level factors. Strikingly, racial and ethnic disparities in COVID-19 illness severity exist, independent of socioeconomic characteristics, which supports the need to further investigate the different levels of racism that contribute to health inequity.(10) Furthermore, non-English language is associated with COVID-19 severity across and within racial/ethnic groups. Efforts should be made to address underlying causes of social inequalities and incorporate such actions into the national COVID-19 response, especially at a time when we have global attention, resources, and focus in hopes to leave this pandemic closer to health equity and not farther from it.(30)

## Data Availability

Data is not publically available at this time as it has not been de-identified and remains on a secure server in accordance with HIPPA policy.

## Acknowledgements

The authors thank Eric Murray and the rest of the MHealth Fairview Information Technology teach of data management support.

## Author Contribution

Concept and design: All authors

Acquisition, analysis, or interpretation of data: Ingraham, Usher, Tignanelli

Drafting of the manuscript: All authors

Critical revision of the manuscript for important intellectual content: All authors

**Appendix Table 1:** ICD10 Codes for Comorbidities. List of ICD 10 codes that were used to classify diagnosis. Abbreviations: T1DM: Type 1 diabetes mellitus, T2DM Type 2 diabetes mellitus, HFpEF: heart failure with preserved ejection fraction, HF: heart failure, CAD: coronary artery disease, VTE: venous thromboembolism, COPD: chronic obstructive lung disease, CKD: chronic kidney disease, ESRD: end stage renal disease, Afib: atrial fibrillation.

| **Comorbidity** | **ICD10 Codes** |
| --- | --- |
| Hypertension | R03.0, I10, I16.0, I11.0, I15.2, I12.9, I11.0 |
| T1DM | E10.21, Z96.41, E10.8, E10.9, E10.10, E10.65, E10.9, E10.641, E10.40, E10.22, E10.69, E10.649, E10.* |
| T2DM | E11.* |
| CAD | I25.* |
| HFpEF | I50.30, I50.31, I50.32, I50.33 |
| Systolic HF | I50.20, I50.21, I50.22, I50.23, I50.40, I50.41, I50.42, I50.43 |
| Any Liver Disease | K70.*, K71.*, K72.*, K73.*, K74.*, K75.*, K76.*, K77.* |
| NAFLD | K76.0 |
| NASH | K75.81, K75.8 |
| Class 2 Obese | Z68.35, Z68.36, Z68.37, Z68.38, Z68.39 |
| Any Obese | E66 |
| Overweight | E66.3, Z68.25, Z68.26, Z68.27, Z68.28, Z68.29 |
| Class 1 Obese | Z68.30, Z68.31, Z68.32, Z68.33, Z68.34 |
| Class 3 Obese | E66.01*, E66.2*, Z68.4*" |
| History of VTE | I27.82, Z86.711, Z86.718, I82* |
| Hypercoagulable state | D68.5*, D68.6* |
| Hypocoagulable state | D68.0*, D68.1*, D68.2*, D68.3*, D68.4*, D68.8*, D68.9*, D69.0*, D69.1*, D69.2*, D69.4*, D69.5*, D69.6*, D69.8*, D69.9*, D66*, D67* |
| Any Heart Failure | I50* |
| COPD | J44*, J41*, J43* |
| Any CKD | N18* |
| Stage 1 CKD | N18.1* |
| Stage 2 CKD | N18.2* |
| Stage 3 CKD | N18.3* |
| Stage 4 CKD | N18.4* |
| Stage 5 CKD | N18.5* |
| ESRD | N18.6*, Z99.2*, T85.631* |
| Afib flutter | I48* |
| Cerebrovascular Disease | G45.*, G46.*, H34.0, I6*.*, I97.81*, I97.82* |
| Sleep Apnea | G47.3* |

**Appendix Table 2:**
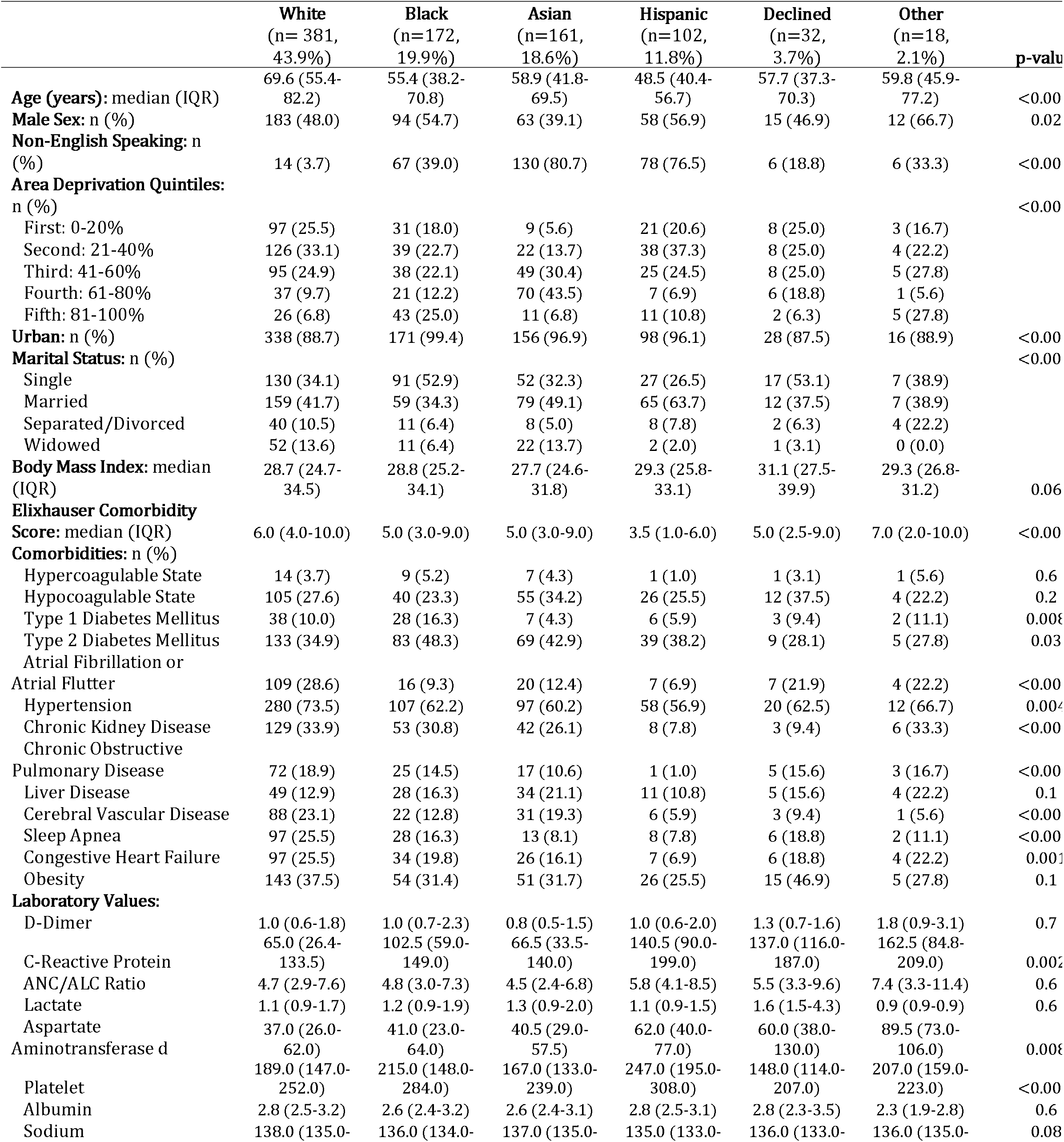
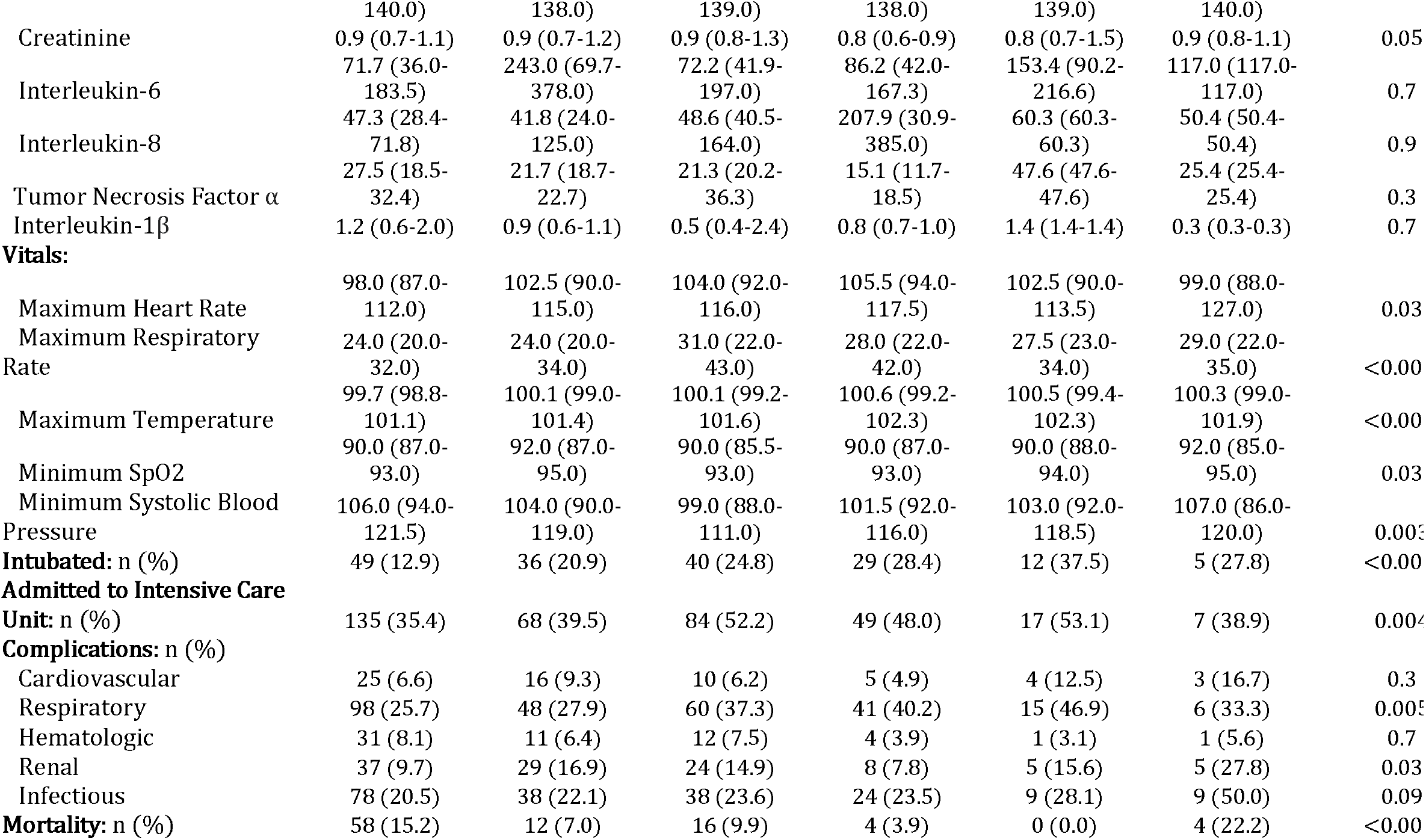
Characteristics of patients admitted to the Hospital by Race/Ethnicity. Univariate analysis of patients hospitalized within 45 days of COVID-19 diagnosis. ADI quintiles represent lowest areas of deprivation (1^st^ quintile) to the highest areas of deprivation (5^th^ quintile).

**Appendix Table 3:** Comparison of lowest to highest level neighborhood deprivation across race/ethnicity. Logistic regression comparing effects of ADI stratified by race (without controlling for the main effects of either race/ethnicity or ADI) on hospitalization within 45 days of diagnosis of COVID-19. ADI quintiles represent lowest areas of deprivation (1^st^ quintile) to the highest areas of deprivation (5^th^ quintile)

|  | **Odds Ratio** | **95% Confidence Interval** | **p-value** |
| --- | --- | --- | --- |
| **Age** | 1.02 | 1.01 – 1.02 | <0.001 |
| **Male Sex** | 1.27 | 1.06 – 1.51 | 0.008 |
| **Race / Area Deprivation Quintiles** |  |  |  |
| White / First: 0-20% | Ref | --- | --- |
| White / Second: 21-40% | 0.92 | 0.66 – 1.29 | 0.63 |
| White / Third: 41-60% | 0.82 | 0.57 – 1.19 | 0.3 |
| White / Fourth: 61-80% | 1.09 | 0.68 – 1.75 | 0.73 |
| White / Fifth: 81-100% | 1.11 | 0.63 – 1.95 | 0.73 |
| Black / First: 0-20% | 1.75 | 1.05 – 2.93 | 0.03 |
| Black / Second: 21-40% | 1.39 | 0.87 – 2.22 | 0.17 |
| Black / Third: 41-60% | 1.32 | 0.8 – 2.16 | 0.28 |
| Black / Fourth: 61-80% | 1.26 | 0.7 – 2.29 | 0.44 |
| Black / Fifth: 81-100% | 1.90 | 1.13 – 3.19 | 0.02 |
| Asian / First: 0-20% | 2.31 | 1.02 – 5.26 | 0.05 |
| Asian / Second: 21-40% | 1.13 | 0.61 – 2.12 | 0.7 |
| Asian / Third: 41-60% | 2.70 | 1.64 – 4.44 | <0.001 |
| Asian / Fourth: 61-80% | 2.42 | 1.5 – 3.91 | <0.001 |
| Asian / Fifth: 81-100% | 4.85 | 1.86 – 12.6 | <0.001 |
| Hispanic / First: 0-20% | 3.68 | 1.97 – 6.88 | <0.001 |
| Hispanic / Second: 21-40% | 3.48 | 2.07 – 5.83 | <0.001 |
| Hispanic / Third: 41-60% | 3.49 | 1.94 – 6.27 | <0.001 |
| Hispanic / Fourth: 61-80% | 2.26 | 0.87 – 5.89 | 0.1 |
| Hispanic / Fifth: 81-100% | 10.14 | 3.85 – 26.74 | <0.001 |
| Decline / First: 0-20% | 2.39 | 1.03 – 5.58 | 0.04 |
| Decline / Second: 21-40% | 1.52 | 0.65 – 3.55 | 0.33 |
| Decline / Third: 41-60% | 1.84 | 0.73 – 4.64 | 0.2 |
| Decline / Fourth: 61-80% | 1.09 | 0.34 – 3.49 | 0.88 |
| Decline / Fifth: 81-100% | 2.31 | 0.51 – 10.38 | 0.28 |
| Other / First: 0-20% | 2.13 | 0.44 – 10.32 | 0.35 |
| Other / Second: 21-40% | 3.02 | 0.83 – 10.98 | 0.09 |
| Other / Third: 41-60% | 2.90 | 0.61 – 13.69 | 0.18 |
| Other / Fourth: 61-80% | 0.45 | 0.06 – 3.62 | 0.45 |
| Other / Fifth: 81-100% | 3.42 | 0.76 – 15.38 | 0.11 |
| **Marital Status** |  |  |  |
| Single | Ref | --- | --- |
| Married | 0.9 | 0.73 – 1.11 | 0.314 |
| Separated | 0.66 | 0.46 – 0.96 | 0.030 |
| Widowed | 0.59 | 0.41 – 0.86 | 0.006 |
| **Non-English Speaking** | 1.87 | 1.46 – 2.4 | <0.001 |
| **Elixhauser Comorbidity Score** | 1.36 | 1.32 – 1.40 | <0.001 |

Conflicts of Interest and Funding Source(s):

1. NIH NHLBI T32HL07741 (NEI)
2. AHRQ R01HS24532 (GBM)
3. AHRQ R01 HS26732 (MGU)
4. AHRQ/PCORI K12HS026379 (CJT)
5. NIH NCATS KL2TR002492, UL1TR002494

Reprints will not be available from the authors. All authors significantly contributed to developing, writing, and revising this manuscript.

The authors have no conflicts of interest to report.

